# The public health impact of fecal microbiota transplantation

**DOI:** 10.1101/2020.04.07.20056952

**Authors:** Lamia Mamoon, Scott W. Olesen

## Abstract

Although fecal microbiota transplantation (FMT) is a recommended, clinically efficacious, and cost-effective treatment for recurrent *Clostridioides difficile* infection (CDI), the scale of FMT use in the United States is unknown. We developed a population-level CDI model and estimated that 48,000 FMTs could be performed annually, preventing 32,000 CDI recurrences.

## INTRODUCTION

*Clostridioides difficile* infection (CDI), the most common hospital-acquired infection in the United States, is a major public health threat, with more than 450,000 US cases in 2017 (1). Aside from the severe diarrhea and other associated health conditions, recurrent CDI can lead to social isolation, humiliation, difficulty keeping a job due to days lost from work and an inability to leave the home, and emotional trauma (2). In 2017, *C. difficile* was estimated as contributing $1 billion to US healthcare costs (3). Approximately 30% of primary CDI patients suffer a recurrence of CDI after standard first-line antibiotic therapy, typically vancomycin or fidaxomicin (4,5). Of patients suffering a first recurrence, approximately 40% recur again (5), and some patients endure months or years in a cycle of disease, antibiotic therapy, remission, and recurrence (6).

Fecal microbiota transplantation (FMT), the transfer of stool from a healthy donor into the gastrointestinal tract of the patient, is a medically recommended treatment option for multiply recurrent CDI (4,7) permitted by the US Food and Drug Administration (8). Evidence from clinical trials suggest that FMT prevents further recurrences in approximately 80% of patients, compared to 20–50% for standard antibiotic therapy (8). Cost effectiveness studies have generally concluded that FMT is superior to standard antibiotic therapy for multiply recurrent CDI (9), and stool banks can relieve individual providers from the logistical complexities of screening donors and manufacturing FMT material (8,10).

Despite the emergence of FMT as a medically recommended and likely cost-effective treatment option for multiply recurrent CDI, the role that FMT does and could potentially play in public health remains poorly quantified. To our knowledge, there are no estimates of the total number of FMTs performed annually in the United States, nor of the number of CDI episodes prevented by use of FMT. To address this gap, we developed a high-level, population-level mathematical model and adapted a recent cost effectiveness model (5) that accounts for patients’ course through multiple CDI recurrences and the role that use of FMT could play in preventing further CDI recurrences. Using this model, we make preliminary estimates of the potential and realized public health benefit of FMT.

## METHODS

The details of the model are in the Supplemental Information. Briefly, the model bins the entire US population as at-risk of CDI, or suffering a primary CDI episode, a first recurrence, a second recurrence, a third recurrence, and so on. Following the approach of Rajasingham *et al*. (5), in the model, some proportion of multiply recurrent CDI patients (i.e., those suffering second or later CDI recurrence, or equivalently a third or later overall CDI episode) are treated with FMT. We call this proportion the “uptake” of FMT. By scaling the uptake from 0% up through 100%, the model estimates the maximum number of FMTs potentially performed annually in the US and the effects of altered uptake on the number of CDI episodes.

Of the model’s 6 input parameters (Supplemental Table 1), 4 were adapted from Rajasingham *et al*. (5) and 2 were drawn from other literature (1,6). The model had 3 outcomes: the annual number of FMTs performed, the proportion of CDI episodes that are multiply recurrent episodes, and the number needed to treat (Supplemental Table 2). In this context, the number needed to treat is the average number of FMTs required to prevent 1 CDI episode.

To account for uncertainty in the input parameters, we drew 1 million bootstrap replicates from distributions around the parameter point estimates (Supplemental Table 1, Supplemental Information). To evaluate the sensitivity of each outcome to the value of each input parameter, we varied each input parameter by 10% around the point estimate and evaluated the fractional change in each outcome.

All simulations and analyses were performed in R (version 3.6.0) (11). The code is available at DOI 10.5281/zenodo.3743343 (https://www.github.com/openbiome/fmt-public-health-model).

## RESULTS

The model estimated that, if FMT were being used to treat all multiply recurrent CDI cases, then 48,000 FMTs (95% CI 34,000 to 62,000) would be performed annually in the US (Supplemental Table 2). The predicted number of FMTs depends strongly on the proportion of multiply recurrent cases treated with FMT (Figure, Supplemental Table 3, Supplemental Figure 1). For example, at 14% coverage, only 10,000 FMTs (95% CI 6,700 to 14,000) are performed. The proportion of CDI episodes that are multiply recurrent (i.e., third or later episodes) was mostly robust to coverage (Supplemental Tables 2 and 3, Supplemental Figure 1). At 0% coverage, the model estimated this proportion at 16% (95% CI 11% to 24%); at 100% coverage, 10% (95% CI 7% to 13%). The estimated number of FMTs required to prevent 1 CDI recurrence was 1.5 (95% CI 0.6 to 4.7) and was very robust to the coverage level (Supplemental Tables 2 and 3, Supplemental Figure 1).

In a sensitivity analysis (Supplemental Table 3, Supplemental Figure 1), the number needed to treat was strongly dependent on the probability of recurrence when using standard therapy: for a 10% increase in the recurrence rate on standard therapy, this outcome declines by 28%. The second strongest dependence in the model was the total number of FMTs on the total number of CDI cases: for a 10% increase in total CDI cases, the number of FMTs increases by 10%.

## DISCUSSION

A population-wide model of the role of FMT in treating CDI recurrences estimated that 10-15% of CDI cases are multiply recurrent. Thus, as many as 48,000 FMTs could be performed in the US annually. The model estimated that, for every 1.5 FMTs performed, 1 CDI episode is prevented. Thus, full use of FMT could prevent as many as 32,000 CDI recurrences (48,000 FMTs / number needed to treat 1.5) every year.

A strength of this model was its evaluation of the population-wide effect of FMT on CDI recurrences. While clinical trials address the merit of FMT for the treatment of individual patients, and cost effectiveness studies address the facility-level economics of FMT, this model estimated the public health impact of this novel therapy. We expect that this model could serve as a scaffold for future refinements and improved estimates.

However, this analysis is subject to important limitations. First, we considered only a single type of CDI, multiply recurrent disease, and a single health outcome, the number of CDI episodes. Emerging evidence suggests that FMT reduces the risk of septicemia in recurrent CDI patients (12) and may reduce mortality when used as treatment for severe and fulminant CDI (13). FMT is also an experimental therapy for dozens of disease indications (8,14). Thus, future studies may need to account for multiple positive outcomes of FMT. Conversely, FMT has important safety risks. Known risks include the transfer of pathogens and complications associated with the delivery procedure. Theoretical risks include the transfer of potentially microbiome-mediated diseases (10,13).

Second, the model estimated FMT’s potential only for preventing CDI recurrences only with respect to the patients treated with FMT. In fact, preventing CDI episodes also has a population-level benefit: fewer CDI episodes in one patient means fewer transmissions and so fewer primary episodes in other individuals. Extending the model to include these population-level effects is possible but will require estimating the total size of the at-risk population and stratifying it by different transmission risks.

Third, the model does not distinguish between patient populations. The estimates for the number needed to treat and the number of CDI episodes prevented are therefore only valid as high-level, population-wide estimates. They may not be applicable for guiding the care of a particular patient or patient population in a particular setting. Stratifying patient populations by disease type, comorbidities, and setting will be important for refined estimates of FMT’s public health benefits as well as its risks.

Finally, we estimated the total possible number of avoided recurrences, but the realized public health impact of FMT depends on the actual number of FMTs performed. OpenBiome, a nonprofit stool bank and likely the largest single FMT material producer in the US, shipped approximately 11,000 FMT preparations in 2019, setting an approximate lower bound for the number of FMTs performed in the US. If only 11,000 FMTs are performed, then FMT coverage could be as low as 15%, and as many as 25,000 preventable CDI recurrences are not prevented (48,000 - 11,000 episodes / 1.5 number needed to treat). We expect that private stool banks and patient-directed procedures account for a substantial number of annual FMTs, but we are not aware of any published estimates of those contributions to the total number of annual FMTs. To add further complexity, there could be simultaneous undertreatment and overtreatment: patients who qualify for FMT may not receive it, and patients for whom FMT is not indicated may be receiving it. Without more transparency around FMT practice, these questions remain topics for speculation only.

Overall, FMT has the potential for enormous positive public health impact, preventing tens of thousands of CDI episodes every year. The FDA’s policy of enforcement discretion, permitting the use of FMT to treat multiply recurrent CDI without an Investigational New Drug application, has enabled widespread use of this therapy (15). Stool banks have also played a key role in expanding safe access to FMT material that can be consistently screened in accordance with international guidelines (10). We encourage other stool banks to publish the number of FMT treatments they manufacture, and hospitals to publish the number of FMTs they perform, so that medical societies and regulators can better understand what barriers remain to widespread, safe access to FMT.

## Data Availability

Only publicly available data were used in this analysis.

https://doi.org/10.5281/zenodo.3743343

## Acknowledgements

We thank Vanessa W. Stevens, Bharat Ramakrishna, and Majdi Osman for helpful comments.

---

**Figure.**
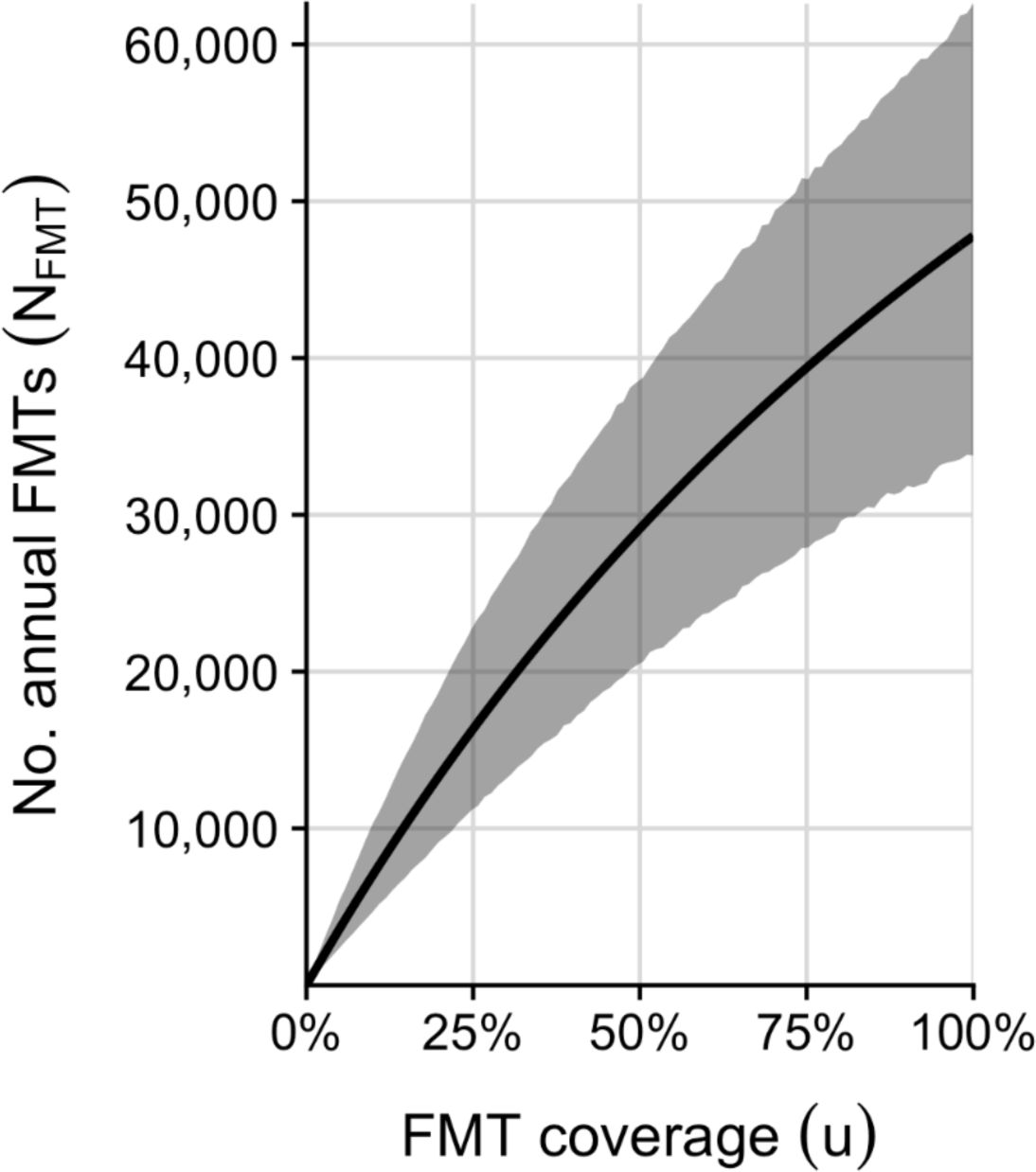
FMT coverage and the annual number of FMTs in the US. Solid line shows the model’s point estimate of the number of FMTs performed annually in the United States as a function of FMT coverage (i.e., the proportion of multiply recurrent CDI cases treated with FMT). Gray region shows 95% quantiles over the 1 million bootstrap variates.

**Supplemental Table 1.** Model parameters.

| Parameter<sup>a</sup> | Point estimate | Range<sup>b</sup> | Statistical distribution | Source<sup>c</sup> |
| --- | --- | --- | --- | --- |
| Probability of first recurrence ($p_1$) | 29% | 25% to 33% | Beta | (5) |
| Probability of second recurrence ($p_2$) | 41% | 29% to 53% | Beta | (5) |
| Probability of third or later recurrence with standard therapy ($p_{abx}$) | 52% | 35% to 70% | Beta | (5) |
| Probability of third or later recurrence with FMT ($p_{FMT}$) | 21% | 17% to 25% | Beta | (5) |
| Annual number of CDI cases ($C$) | 462,100 | 428,600 to 495,600 | Normal | (1) |
| Maximum number of CDI episodes ($E$) | 15 | 10 to 20 | Uniform | (6) |

a: Mathematical notation of parameters is as in the Supplemental Information.
b: For beta- and normal-distributed variables, the reported range is the 95% confidence interval. For the uniform-distributed variable, it is the minimum and maximum values.
c: See the Supplemental Information for how the values reported in the literature were adapted to the statistical distributions. The range for $E$ was chosen as one-third range around the point estimate.

**Supplemental Table 2.** Model outcomes.

| Parameter<sup>a</sup> | FMT coverage ($u$) | Point estimate (95% CI) |
| --- | --- | --- |
| No. annual FMTs ($N_{\text{FMT}}$) | 0% | 0 |
|  | 50% | 29,000 (20,000 to 39,000) |
|  | 100% | 48,000 (34,000 to 62,000) |
| Proportion of CDI episodes that are third or later episodes ($f_{\geq 3}$) | 0% | 16% (11% to 24%) |
|  | 50% | 13% (9% to 17%) |
|  | 100% | 10% (7% to 13%) |
| Number needed to treat, for FMT vs. standard therapy (NNT) | 0% | 1.5 (0.6 to 4.7) |
|  | 50% | 1.5 (0.6 to 4.7) |
|  | 100% | 1.5 (0.6 to 4.7) |

a: Mathematical notation as used in the Supplemental Information.

**Supplemental Table 3.** Sensitivity analysis. Only combinations of parameters and outcomes with sensitivities greater than 0.1% are shown.

| Parameter<sup>a</sup> | Outcome<sup>b</sup> | Sensitivity<sup>c</sup> |
| --- | --- | --- |
| $p_{\text{abx}}$ | NNT | −28% |
| $u$ | $f_{\geq 3}$ | −2.2% |
| $p_{\text{FMT}}$ | $N_{\text{FMT}}, f_{\geq 3}$ | 1.4% |
| $p_{\text{abx}}$ | $N_{\text{FMT}}, f_{\geq 3}$ | 3.6% |
| $p_{\text{FMT}}$ | NNT | 6.4% |
| $p_1$ | $N_{\text{FMT}}, f_{\geq 3}$ | 6.8% |
| $u$ | $N_{\text{FMT}}$ | 7.8% |
| $p_2$ | $N_{\text{FMT}}, f_{\geq 3}$ | 8.7% |
| $C.$ | $N_{\text{FMT}}$ | 10% |

a: Parameters as annotated in Supplemental Table 1
b: Outcomes as annotated in Supplemental Table 2
c: Percent change in outcome per 10% increase in parameter

**Supplemental Figure 1.**
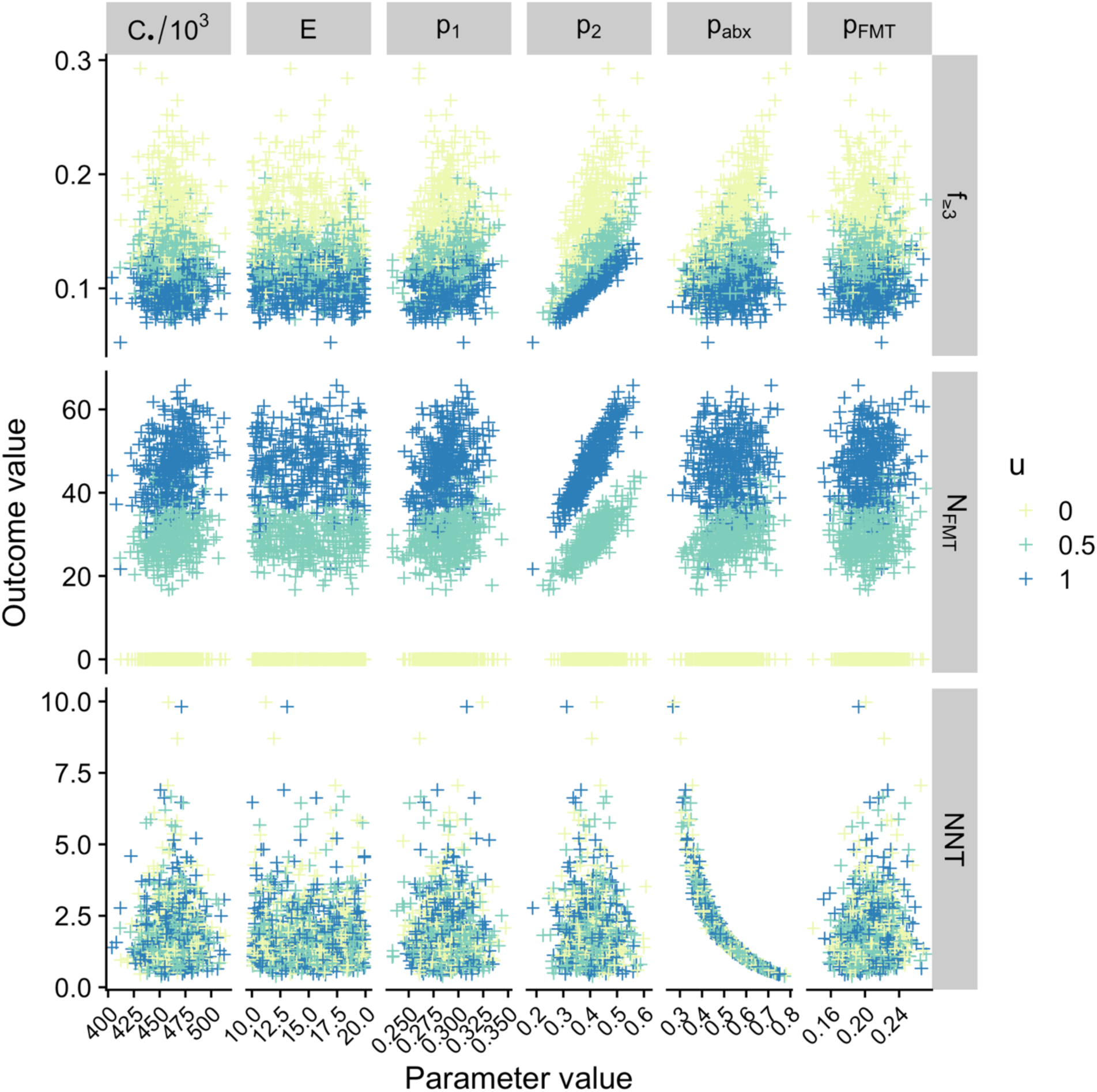
Input parameter values (horizontal axis, top labels) and output outcome values (vertical axis, right labels) for three values of FMT coverage *u* (colors). For visual clarity, the 0.3% of bootstrap replicates with a number needed to treat outside of 0 to 10 were excluded, and a random subset of 1,000 of those qualifying bootstraps is shown.

